# Novel Entropy-Based Framework for Quantifying Dynamic Epistemic Uncertainty in Clinical Medicine

**DOI:** 10.64898/2026.08.27.26361497

**Authors:** Yuichiro Yano, Eigo Shintani, Shunya Arita, Rei Ashine, Naoaki Iinuma, Hirotake Mori, Kazutoshi Fujibayashi, Yosuke Yamada, Mizue Saita, Naoki Nakashima, Hiroshi Itoh, Masaomi Nangaku, Mizuki Ohashi, Hiroyuki Daida, Hajime Arai, Toshio Naito

**Affiliations:** Department of General Medicine, Juntendo University Faculty of Medicine, Tokyo, Japan; Artificial Intelligence Incubation Farm, Juntendo University Faculty of Medicine, Tokyo, Japan; SoftBank Corp., Tokyo, Japan; Medical Information Center, Kyushu University Hospital, Japan; Center for Preventive Medicine, Keio University School of Medicine, Tokyo, Japan; Division of Nephrology and Endocrinology, The University of Tokyo Graduate School of Medicine, Tokyo, Japan; Department of Cardiovascular Biology and Medicine, Juntendo University Graduate School of Medicine, Tokyo, Japan; Department of Neurosurgery, Juntendo University, Tokyo, Japan

## Abstract

The widespread adoption of clinical large language models (LLMs) introduces significant risks of automation bias, premature closure, and clinician deskilling. Current interpretability paradigms, including latent space trajectories, Concept Activation Vectors, and Concept Bottleneck Models, suffer from topological stagnation, metric distortion, and epistemic occlusion, frequently masking intermediate diagnostic uncertainty behind falsely confident outputs. To address these structural vulnerabilities, this paper introduces a novel closed-loop, multi-agent framework designed to quantify and visualize dynamic epistemic uncertainty in clinical LLM reasoning. By coupling predictive Shannon entropy with non-linear Isometric Feature Mapping (ISOMAP), the architecture projects high-dimensional inference state vectors onto a calibrated two-dimensional latent space, thereby assigning a quantifiable thermodynamic energy state to the reasoning path to track diagnostic velocity, cognitive momentum, and trajectory efficiency across sequential diagnostic rounds.

Pilot validation across representative emergency medicine scenarios demonstrated distinct topological and information-theoretic behaviors: unconfounded cases (cerebellar infarction) exhibited smooth geodesic progression toward the ground truth alongside monotonic Shannon entropy decay from 2.15 to 1.74; noisy environments with ambiguous findings (spontaneous pneumothorax) suffered from trajectory wandering, local minimum traps, and high sustained entropy (~2.41) due to insufficient repulsive weighting for negative evidence; and triage-conflicted cases (acute cholangitis) achieved precise geometric proximity to the true node but experienced top-1 rank stagnation because the model conflated acute severity triage (sepsis) with anatomical etiology. By rendering machine hesitation and cognitive divergence visually auditable before final diagnostic crystallization, this geometric-information framework enables dynamic trust calibration and human-AI co-regulation at the point of care while establishing a clear mathematical foundation for future architectural interventions, such as dual-channel safety decoupling and non-linear repulsive weighting. Moving forward, validating these architectural enhancements across large-scale electronic health record databases and prospective clinical trials will be essential to realize its full clinical utility, establishing a foundational blueprint for safe, transparent, and cognitively synergistic AI integration in future medical practice.

By rendering the LLM’s reasoning process visually auditable, this framework lays the groundwork for capturing and externalizing the clinician’s own cognitive patterns within the AI, forming a coupled system. This enables the explicit visualization of cognitive gaps between physician hypotheses and AI inferences, transforming the interaction from simple answer-checking into a dynamic learning process for both human and machine that prevents diagnostic oversight. Ultimately, because the responsibility for final clinical decision-making remains with the human practitioner, this framework serves as a vital decision-support mechanism. Moving forward, validating these architectural enhancements across large-scale electronic health record databases and prospective clinical trials will be essential to realize its full clinical utility, establishing a foundational blueprint for safe, transparent, and cognitively synergistic AI integration in future medical practice.

## 1. Introduction

The rapid proliferation of clinical large language models (LLMs) and generative artificial intelligence (GenAI) has fundamentally revolutionized diagnostic precision and workflow efficiency in clinical medicine (1-3). However, this widespread adoption introduces severe cognitive biases and an insidious deskilling effect that threatens the core of clinical reasoning (4,5). Recent empirical data highlights a profound anxiety within the medical community: 77% of physicians and 70% of nurses express deep concern over the atrophy of their clinical skills due to over-reliance on automated systems (6). Two prominent cognitive pathologies arising from this shift are automation bias—the uncritical acceptance of system outputs—and premature closure, where clinicians prematurely crystallize a diagnostic hypothesis and exclude critical alternatives (7-9).

This erosion of human expertise is an observable clinical reality. Evidence demonstrates that continuous exposure to diagnostic AI can render clinicians less motivated, less focused, and less accountable during independent decision-making (10,11). The speed of this cognitive degradation was starkly illustrated in a recent randomized study of expert endoscopists: after exposure to a real-time AI lesion-flagging system, physicians’ independent adenoma detection rates (ADR) plummeted from a baseline of 28.4% to 22.4% on days when the AI was unavailable (12).

This alarming trend reveals a dangerous paradox in modern medicine: clinicians may perform at a deceptively high level by temporarily borrowing skills from AI, yet they fail to internalize or retain that expertise. Unlike historical technologies that automated manual labor, GenAI uniquely automates the core cognitive faculties of thinking and interpretation. Mitigating these vulnerabilities requires a profound shift in workflow design, balancing algorithmic convenience with the absolute necessity of mindful vigilance at the point of care.

## 2. Cognitive and Structural Vulnerabilities in Human-AI Co-Regulation

### 2.1 Cognitive Mechanics of Premature Closure and Overreliance

Automation bias represents a critical cognitive propensity where human operators prioritize automated system outputs over their own clinical judgment, even when faced with contradictory, objective empirical evidence (7-9). This cognitive vulnerability has become acutely pronounced with the introduction of highly autonomous GenAI. As established by Logg et al., human decision-makers exhibit a marked tendency toward *algorithmic appreciation*, systematically overvaluing algorithmic advice over human counsel when tasked with complex, objective operations (13). In medicine—a highly specialized domain inherently fraught with epistemic uncertainty—this appreciation escalates dangerously. When acute time constraints and cognitive fatigue from demanding shift schedules are superimposed, clinicians are increasingly predisposed to perceive automated systems as infallible or oracular. Consequently, they abandon autonomous clinical hypothesis generation, triggering a cognitive lock-in that prematurely converges on the AI’s initial recommendation (7, 14,15).

This dynamic closely mirrors historical clinical errors documented in traditional computer-aided detection systems, where radiologists routinely over-relied on automated prompts and overlooked visible abnormalities missed by the software (16,17). Reflecting this precedent, modern healthcare environments are witnessing the structural manifestation of omission errors: clinicians uncritically accept an AI’s non-detection output, skip meticulous independent readings, and consequently overlook hidden or subtle lesions.

When clinicians habitually treat AI recommendations as default truths and neglect active, independent verification, cognitive complacency becomes deeply entrenched. To quantitatively assess this overreliance, contemporary medical AI benchmarks utilize metrics such as the switch fraction—the rate at which a user overturns an initially correct clinical judgment to align with an erroneous AI recommendation—and the overall probability of uncritical acceptance of false recommendations, termed accept-on-wrong (18,19). Strikingly, individuals with low AI literacy are disproportionately susceptible to these cognitive biases, largely due to a fundamental deficit in recognizing the inherent limitations of automated systems. Lacking a granular understanding of AI’s strict reliance on preprogrammed algorithms and statistical pattern recognition rather than genuine comprehension, they often fail to grasp its absolute inability to navigate human emotions, social nuances, and moral complexities (20). This deficiency stifles the necessary epistemic skepticism required at the point of care, predisposing low-literacy users to overtrust the system’s capacity to evaluate subjective clinical contexts (21).

### 2.2 Classification of Dependence in Clinical Decision-Making

Dissecting the classification of dependence in clinical decision-making reveals that system safety within the human-AI collaborative dynamic is governed neither by the average accuracy of the model nor by the generic trust of the user. Instead, it is strictly dictated by the precision of *trust calibration*: the physician’s dynamic capacity to appropriately accept or reject system outputs based on the contextual correctness of the AI across highly heterogeneous clinical scenarios. Mitigating these cognitive vulnerabilities therefore requires moving beyond static, outcome-based metrics, highlighting an urgent prerequisite to visualize and audit the precise trajectory, direction, and depth of clinical reasoning during human-AI interactions.

## 3. Current Paradigms in Clinical Reasoning Visualization

To contextualize the necessity of advanced trust calibration, this section reviews the three dominant geometric and structural paradigms currently utilized to visualize and audit clinical reasoning in artificial intelligence systems.

### 3.1 Latent Space Trajectories on the Medical Manifold

The first paradigm conceptualizes highly heterogeneous, multimodal clinical data streams—such as genomics, electronic health record (EHR) time-series, and high-resolution imaging—as high-dimensional realizations constrained by an underlying, integrative low-dimensional medical manifold (22-24). Utilizing deep neural encoders, distinct modality inputs x_*m*_ (for *m* ∈ *M*) are compressed into a shared *d*-dimensional latent space *Z*, yielding a unified embedding vector *z*_*t*_:

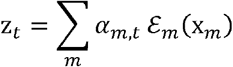

where ℰ_*m*_ represents the latent representation map of modality *m*, and *α*_*m,t*_ denotes the dynamic cross-modal attention weight at time *t*. Within this continuous geometry, a patient’s diagnostic state is represented as a dynamic trajectory z(*t*), while therapeutic interventions are formalized as a velocity vector ż(*t*) governed by a transition operator *G* and a contextual clinical intervention vector *u*(*t*):

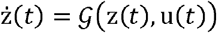

By visually inspecting the proximity of z(*t*) to historical case clusters and the geometric topology of diagnostic boundaries, clinicians can intuitively audit the model’s evidence-integration process.

However, down-projecting this high-dimensional clinical knowledge space onto a localized two-dimensional manifold for visual trajectory tracking inevitably introduces three critical geometric pathologies. First, metric and scale distortion corrupts both global and local distance metrics during non-linear dimensionality reduction. Consequently, an extensive spatial translation of a trajectory vector between clusters representing distinct conditions, such as myocardial infarction and aortic dissection, becomes visually ambiguous, making it impossible to determine based purely on geometric topology whether the displacement reflects a substantial, evidence-driven paradigm shift or merely low-magnitude stochastic noise inherent to the latent space compression. Second, path morphological equifinality arises because disparate clinical journeys can culminate in identical diagnostic coordinates. For instance, while two clinicians may reach the same final domain for acute appendicitis, their geometric trajectories often differ fundamentally, meaning that purely spatial parameters cannot establish an objective baseline to discern whether a direct, linear path represents refined diagnostic mastery or a convoluted, high-curvature trajectory signifies an ungrounded novice who arrived at the correct diagnosis through cognitive wandering. Third, topological boundary stagnation occurs as a trajectory approaches a critical saddle point or decision boundary separating phenotypically similar pathologies. In these regions, spatial velocity ‖ż(*t*)‖ drastically decays, yet visual inspection alone cannot determine whether this kinetic arrest reflects a healthy clinical deliberation involving balanced information seeking or an anomalous software freeze predictive of an impending hallucination.

#### Thermodynamic Resolution via Geometric-Information Coupling

To mitigate these intrinsic topological limitations, we introduce an information-theoretic framework that superimposes an orthogonal thermodynamic axis—predictive Shannon entropy *H*(*t)*—onto the structural geometry of the latent trajectory. This thermodynamic coupling imbues the passive geometric lines with a quantifiable cognitive energy state, thereby resolving the three structural pathologies.

Principally, this integration exposes the operational dynamics of the clinical reasoning process by formalizing a dynamic cognitive momentum indicator, computed as the product of geometric velocity and information certainty. This formulation enables a definitive classification of kinetic states based on the interplay between entropy and velocity. For instance, a regime characterized by low entropy (*H*→ 0) and high velocity (‖ż(*t*)‖0) indicates that the system is actively accelerating through the hypothesis space with deterministic clinical confidence. Conversely, the co-occurrence of high entropy (*H* → *H*_max_) and velocity stagnation (‖ż(*t*)‖ 0) signifies that the system has safely applied a cognitive brake at a critical topological bifurcation to execute a deliberate examination of conflicting clinical evidence. Finally, a combination of high entropy (*H* → *H*_max_) and high velocity (‖ż(*t*)‖0) betrays a pathological state of unanchored divergence, where the system oscillates erratically across diagnostic boundaries due to acute epistemic confusion and instability.

Furthermore, this framework mathematically formalizes a Trajectory Efficiency Score (*η*) by integrating the continuous rate of entropy decay over the entire reasoning path *P*:

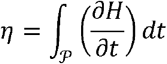

Under an expert diagnostic regime, *η* reveals a highly efficient, monotonic decay of thermodynamic energy as clinical information is sequentially internalized, driving the entropy cleanly toward zero. In contrast, a novice trajectory displays high information-theoretic turbulence, marked by erratic entropy oscillations that capture the precise magnitude of wasted cognitive energy and structural reasoning inefficiencies.

To operationalize this geometric-information coupling, mapping high-dimensional reasoning state vectors, high-dimensional reasoning state vectors, which represent probability distributions across candidate hypotheses, are mapped onto a calibrated two-dimensional latent space using Isometric Feature Mapping (ISOMAP). By anchoring disease nodes via barycentric embeddings, this approach visually and mathematically exposes whether an agent’s reasoning trajectory cleanly aligns with the true diagnosis or becomes trapped in confounding attractors.

### 3.2 Post-Hoc Semantic Alignment via Concept Activation Vectors (CAVs)

While manifold trajectories track holistic state shifts, Concept Activation Vectors (CAVs) illuminate the specific semantic weights of high-level, abstract medical concepts (e.g., “nuclear atypia,” “exudates”) that traditional pixel-level saliency maps fail to capture (25-27). In the Testing with CAV (TCAV) framework, a linear classifier is trained on intermediate layer *ℓ* activations to separate positive concept exemplars from negative controls. The CAV 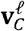 is defined as the unit normal vector to this separating hyperplane. The conceptual sensitivity 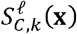 of an input **x** for a target diagnostic class *k* is subsequently calculated as the directional derivative of the decision bottleneck layer 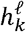:

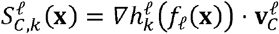

where *f*_*ℓ*_(**X**) represents the activation vector at layer *ℓ*, and 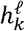 maps this representation to the logit space of class *k*. Recent advancements, such as *σ*-TCAV, utilize parametric smoothing via generalized sigmoids under a stochastic Bayesian formulation, successfully reducing empirical variance by approximately 50% to mitigate the statistical instability inherent in classic multi-layer CAV ensembles.

However, despite its capacity for semantic grounding, the CAV framework exhibits two critical vulnerabilities when auditing dynamic clinical reasoning: an inability to account for out-of-distribution (OOD) epistemic uncertainty, and a fundamental conflation of directional alignment with predictive confidence. First, because CAVs require human experts to define exemplar sets *a priori*, they suffer from semantic blindness when a model encounters unanticipated clinical anomalies or latent, OOD phenotypes that fall outside these pre-configured conceptual boundaries; under these conditions, CAV metrics often remain orthogonal to the anomalous features or yield uninterpretable outputs. Second, analyzing the directional geometry of 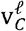 alone cannot differentiate whether the model is propagating along a specific disease trajectory with deterministic certainty, or merely tracing a high-variance, pseudo-random path driven by stochastic dispersion.

To overcome these geometric blind spots, we introduce an information-theoretic complementarity framework leveraging predictive and attention entropy to directly quantify the magnitude of the model’s internal epistemic uncertainty. This dual approach yields distinct diagnostic advantages across two pathological reasoning extremes. First, in detecting pathological divergence (stochastic dispersion), when a LLM encounters a dense differential diagnosis and disperses its probability mass across mutually exclusive clinical hypotheses, traditional CAVs risk misinterpreting this high-dimensional alignment as a structured, deliberate evaluation of rare conditions. In contrast, computing the predictive entropy of the next-token probability distribution, *H* (*P*) = − ∑_*i*_ *P* (*x*_*i*_) log_2_ *P* (*x*_*i*_), reveals a sharp entropy surge. This quantitative signal alerts the clinician that the model’s predictive distribution has degraded into high-variance stochasticity, yielding ungrounded clinical trajectories. Second, in diagnosing premature convergence (epistemic closure), during premature clinical closure—where a model high-confidently crystallizes a benign diagnosis (e.g., common viral pharyngitis) early in the clinical sequence despite insufficient diagnostic evidence—a traditional CAV merely registers a high sensitivity score for that specific concept, failing to capture the underlying systemic flaw. Our information-theoretic audit flags this anomaly via the premature collapse of both predictive and attention entropy toward zero. Identifying this asymptotic decay prior to adequate contextual ingestion provides a rigorous mathematical trigger, warning clinicians of ungrounded overconfidence and the systematic exclusion of valid alternative pathways.

### 3.3 Intrinsic Bottlenecks and Interventional Editing via CBMs

Moving beyond post-hoc explanations, Concept Bottleneck Models (CBMs) intrinsically restructure the forward pass by embedding a human-interpretable concept layer directly into the neural bottleneck. To prevent the model from exploiting spurious correlations that violate established clinical practice guidelines, a perturbation-based alignment loss (ℒ_align_) is introduced during optimization:

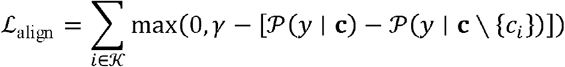

where *K* represents the index set of critical clinical concepts, **c**\{*c*_*i*_} denotes the concept vector with the *i*-th concept masked, and *γ* is a pre-specified probability drop threshold enforced when a critical concept is absent [28, 29].

#### 3.3.1 Epistemic Occlusion and the Limitations of Standard CBMs

Despite these architectural advantages, standard CBM formulations suffer from a critical diagnostic limitation: the occlusion of epistemic uncertainty during downstream task projection. First, because CBMs propagate information via a rigid, unidirectional pipeline (**x**→**c** → *y*), a highly unresolved, divergent concept state—wherein multiple conflicting clinical attributes hover near an equivocal probability of 0.5—is frequently compressed and flattened during final logit aggregation. Consequently, the downstream task predictor may output a seemingly definitive diagnosis (e.g., allocating a dominant probability mass to a single disease), leaving the clinician entirely blind to the severe hesitation and mathematical divergence latent within the intermediate concept space.

Second, conventional CBM task predictors are predominantly optimized under a mutually exclusive, multi-class classification paradigm utilizing a standard Softmax activation layer. This mathematical constraint forces the model to distribute a single probability mass across all targets, effectively conflating genuine diagnostic ambiguity (where multiple distinct, competing diagnoses simultaneously co-exist) with uniform, low-confidence background noise.

## 4. Methods: Pilot Study Design and Framework Architecture

The AI’s sequential clinical reasoning system proposed in this study is a closed-loop, multi-agent collaborative framework designed to conceptually emulate the decision-making process of expert physicians in emergency clinical settings. Unlike conventional static language model applications that output a single judgment from a single input, this AI’s system comprises independent modules for diagnostic hypothesis generation, execution of clinical judgments, simulation of the diagnostic environment, update of ranking of diagnostic hypotheses, and evaluation of the quality of these clinical judgments. These modules operate autonomously according to their dedicated roles, forming a recursive flow via a feedback mechanism to determine hypothesis rankings. In the followings, we call “round” as one cycle of such clinical judgment flow.

The clinical judgment module does not merely choose the “test that most reduces uncertainty,” but introduces a quality score to resolve the conflicting medical constraints of ensuring clinical safety and minimizing patient burden. Furthermore, to prevent fluctuations in diagnostic hypothesis ranking, which means the sort of hypothesis according to its probability given from AI’s system, under the noisy environments (this is the environment containing indeterminable/borderline pseudo-test result in simulator) akin to real clinical practice, a control filter is implemented to regulate probability updating and ranking stability.

The clinical judgment evaluation module maps the execution logs to a quantified score of quality of process. Furthermore, to visualize the thinking process of the AI’s system over time, a high-dimensional inference state vector is constructed from the inference probability distribution. ISOMAP, a non-linear dimensionality reduction algorithm that preserves geodesic distances between data points, is applied in order to map the inference state at each diagnostic step onto a two-dimensional plane (see also section 5).

Moving beyond single-prompt LLM execution, the above framework decouples reasoning execution, environment simulation, belief updating, and process auditing across four autonomous agents. Furthermore, by formulating test selection as a multi-objective optimization problem that balances entropy reduction, contrastive updates, safety constraints, and patient burden, and by enforcing threshold-based order-locking mechanisms, our approach eliminates cognitive wandering and achieves robust, deterministic convergence toward the true diagnosis.

### 4.1. Information-Theoretic Resolution via Concept Entropy Mapping

To explicitly audit whether a differential diagnosis of our proposed AI’s system is robustly converging or pathologically diverging, it is essential to inject information-theoretic uncertainty estimation into the concept bottleneck topology. Instead of relying solely on task-level logits, we propose quantifying the structural entropy of the intermediate concept layer. Given a computed concept probability vector **c** = [*c*_1_,*c*_2_,…,*c*_*m*_)^*T*^, where each element *c*_*j*_ ∈ [0,1] reflects the calibrated predicted presence of an individual clinical predicate from AI’s system, the total Concept Information Entropy *H* (**c**) is formalized using the sum of Shannon entropies:

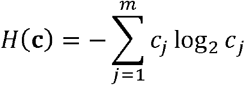

Under a convergent diagnostic regime, only pathognomonic clinical hypothesis is close to 1.0 while others drop to 0.0, strictly minimizing *H*(**c**) and indicating a well-circumscribed, highly deterministic diagnoses can be realized. Conversely, during clinical divergence, incompatible or poorly generalized concepts hover within a maximum-uncertainty plateau (*c*_*j*_ ≈ 0.5), driving *H* (**c**) to its theoretical maximum and structurally signaling a fragmented inference path. Real-time visualization of this dynamic concept-entropy metric effectively contextualizes the AI’s “internal confusion” for the physician before final hypothesis crystallization.

## 5. Results: Pilot Validation and Geometric Trajectory Analysis

To intuitively validate the clinical efficacy of our framework, we focus on three representative scenarios that most symbolically describe the clinical reasoning process generated using GPT-5.3 Instant [OpenAI, 2026]: typical convergence case, wandering and mis-convergence case, and stagnation case. The decision-making trajectories are visualized by using ISOMAP algorithm. The inference history logs generated from initial direct-instruction LLM configuration in the three representative cases are comparable for those clinical efficacies.

ISOMAP algorithm can present the probability distribution of hypotheses mapped onto a calibrated two-dimensional latent coordinate space defined by Dim 1 (horizontal axis) and Dim 2 (vertical axis) utilizing non-linear dimensionality reduction. Here the inference state vector is given from the probability of hypotheses. Spatial proximity within two state vectors directly reflects similarity of two probability distributions.

In the below, the visual annotation schemas used in our results are represented:

### Averaged Position of Hypothesis

We represent the weighted averaged coordinates corresponding to individual hypotheses,

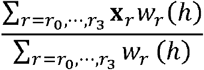

where **x**_*r*_ denotes the vector of ISOMAP coordinate at round *r*, and *w*_*r*_(*h*) denotes the weight value which consists of the probability of hypothesis *h* ∈ [*H*1,*H*2,…] at round *r*. This represents the reference position for the comparative hypothesis with ground truth (see below).

### Ground Truth

This is the averaged position of hypothesis corresponding to the true definitive diagnosis.

### Trajectory from *r*_O_ **to** *r*_3_

The trace of evaluated ISOMAP coordinate of the AI’s inference state vector across the diagnostic rounds of closed-loop step. This is regarded as the clinical reasoning trajectory.

### Point of Top-1 Wrong Answer

we highlight the inference state vector of which the AI agent assigns its highest probability rank (Top-1 prediction) to an incorrect hypothesis.

In the following, the reasoning trajectory originated from the bottom-right baseline coordinate (*r*_O_, regarding the initial patient intake) is a sequential transition line through subsequent diagnostic rounds (*r*_1_ → *r*_2_ → *r*_3_, regarding the progressive test execution and belief updating). The geometric morphology and directional velocity of this trajectory provide deterministic visual audit of the AI’s internal cognitive state. Our representative scenario has three different transitions achieved or not achieved correct diagnosis (close to ground truth) and reasons of such issues:

### Direct Geodesic Progression

Smooth, linear movement along the shortest path toward the target node signifies healthy, evidence-driven evidence integration and confident convergence.

### Geometric Deviation and Local Minimum Traps

Non-linear wandering or deflection toward non-target clusters exposes cognitive distraction by clinical pseudo-noise or an inability to process negative evidence.

### Spatial Convergence with Top-1 Error Stagnation

Trajectories that physically reach the immediate vicinity of the ground-truth node yet remain enclosed by Top-1 error squares indicate structural reasoning bottlenecks, such as the conflation of acute severity triage with anatomical etiology.

### 5.1. Case A: Cerebellar Infarction in a Noisy Environment

- **Patient Profile:** 68-year-old male
- **Chief Complaint:** Sudden onset of vertigo, vomiting, and gait disturbance
- **History of Present Illness:** Onset of continuous rotational vertigo and vomiting upon awakening in the morning. An antiemetic was prescribed at a local clinic without symptom relief, and he was transported via emergency medical services due to an inability to walk. The rotational vertigo was continuous and mildly exacerbated by changes in head position, yet persisted at rest. No overt diplopia was noted. The patient exhibited severe difficulty maintaining a standing posture. Tinnitus was absent. Mild rhinorrhea was reported a few days prior.
- **Past Medical History:** Hypertension, paroxysmal atrial fibrillation
- **Medications:** Antihypertensives; oral anticoagulant therapy self-discontinued
- **Allergies / Social History:** No known drug allergies; positive smoking history
- **Family History:** Father with cerebral infarction
- **Vital Signs:** Body Temperature: 36.8°C, Heart Rate: 88 bpm, Blood Pressure: 164/92 mmHg, Respiratory Rate: 18 breaths/min, SpO_2_: 98% (room air), Japan Coma Scale (JCS): 0

As a control case where inference proceeded normally and reached the correct answer, the inference trajectory of cerebellar infarction in a noisy environment is shown in **Figure 1**. The patient is assumed to be transported to the emergency department presenting continuous rotational vertigo, vomiting, and gait disturbance. The inference state at the initial state *r*_O_ immediately following the medical interview (the lower-right point in the latent space) is on the position furthest from the ground-truth hypothesis, cerebellar infarction.

**Figure 1:**
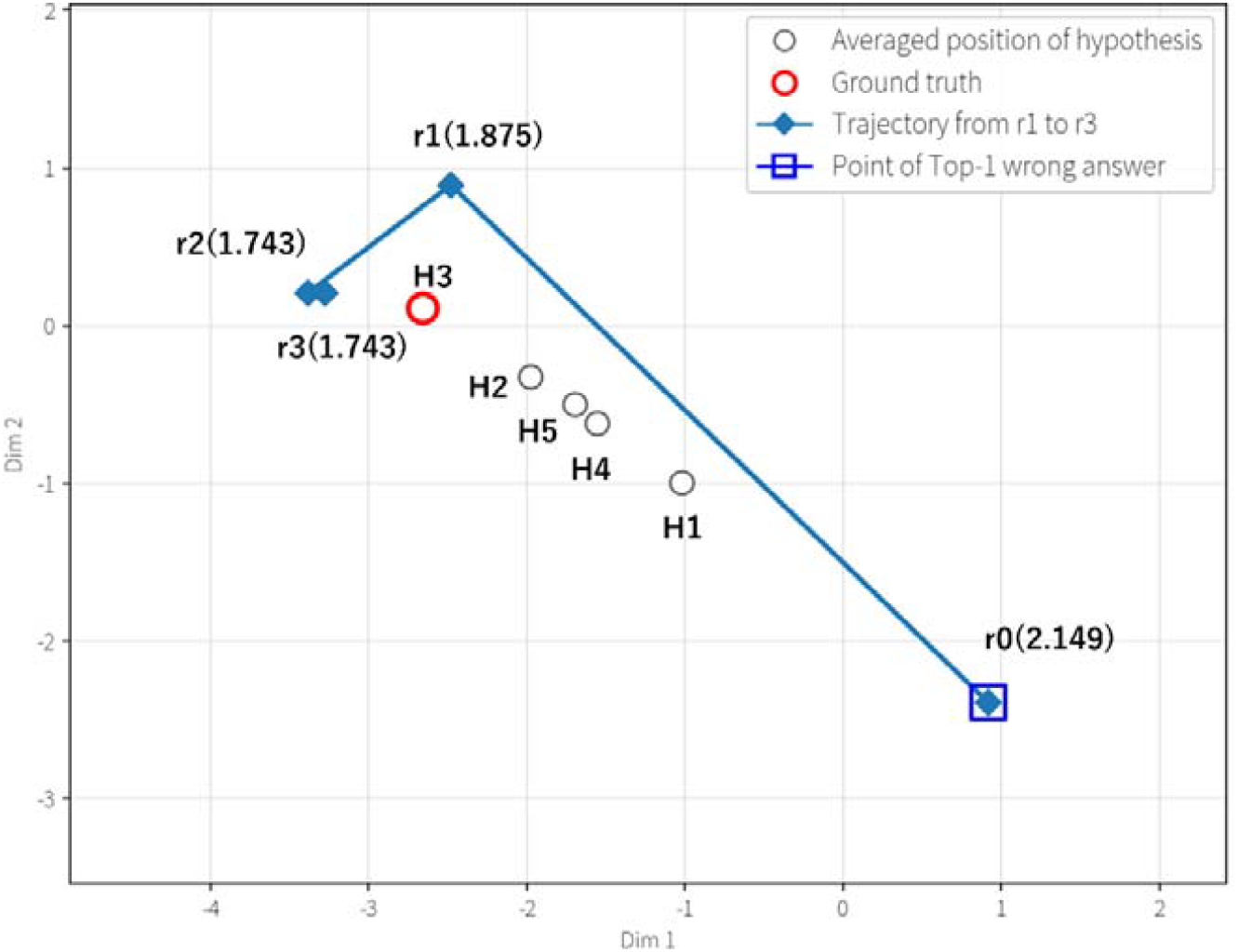
ISOMAP trajectory (filled diamond) for the ground truth disease, cerebellar infarction at each round (showing “”). Converged near the ground truth disease far from the initial value. Here, the white circles represent as the reference locations of hypothesis, where H1 indicates vestibular neuritis, H2 indicates brainstem infarction, H3 indicates cerebellar infarction which consists with ground truth (red circle), and H4 indicates Benign Paroxysmal Positional Vertigo. The numerical value in the parentheses of “*r*” denotes the entropy after each round.

In the first round (*r*_1_), the AI’s system proposes the high-information tests (such as physical assessments and neuroimaging) required for the differential diagnosis of vertigo (central vs. peripheral), and the diagnostic environment simulator returns pseudo-test results for these candidate interventions. After the update of probability distribution according to this clinical judgment, the inference state vector (blue diamond) jumped to the upper-left region near a position of ground truth.

Subsequently, from the second round (*r*_2_) update to the final third round (*r*_3_), the inference plot clearly bypasses the competing peripheral and central nodes, such as vestibular neuritis (H1) and brainstem infarction (H2), and proceeds smoothly along the shortest geodesic line. In the final round (*r*_3_), the inference state vector converges onto the barycenter of cerebellar infarction (H3) which is a target coordinate designating the ground truth hypothesis. This suggests that in standard clinical testing, the AI’s system possesses the capability to converge seamlessly without wandering from an initial value far removed from the correct hypothesis.

As the inference state vector is updated at each round, Shannon entropy is smoothly decreased from initial round value 2.15 to final round one 1.74 (**Table 1 and Figure 1**) and its value is seen to being stable at final round. This indicates that the convergence of ISOMAP inference state vector onto the ground-truth, closing geometric position between neighbor points, is also strongly relevant to the variance of Shannon entropy.

**Table 1:** Showing the probability of each hypothesis at initial () and after a round from the first () to third () with those entropy values, given from AI’s system in case A.

| Round |  |  |  |  |  | Entropy |
| --- | --- | --- | --- | --- | --- | --- |
|  | 0.333 | 0.2667 | 0.2 | 0.1333 | 0.0667 | 2.149 |
|  | 0.12 | 0.25 | 0.5 | 0.08 | 0.05 | 1.875 |
|  | 0.1 | 0.23 | 0.56 | 0.07 | 0.04 | 1.743 |
|  | 0.1 | 0.23 | 0.56 | 0.07 | 0.04 | 1.743 |

### 5.2. Case B: Spontaneous Pneumothorax in a Noisy Environment

- **Patient Profile:** 31-year-old male
- **Chief Complaint:** Chest pain and shortness of breath
- **History of Present Illness:** Onset of chest discomfort and shortness of breath in the morning, which gradually worsened over time. The patient noted mild pain upon inspiration. No fever or cough was present. Mild discomfort in the left lower leg was reported. Severe unilateral chest pain was not clearly identified.
- **Past Medical History:** Unremarkable
- **Medications / Allergies:** No regular medications; no known drug allergies
- **Social / Family History:** Current smoker (10 cigarettes/day); history of a long-distance flight 2 days prior. Positive family history of thrombosis.
- **Vital Signs:** Heart Rate: 116 bpm, SpO_2_ : 92% (room air)

As a representative example where an inference error occurred under indeterminate/borderline findings (a pseudo-noise environment), the decision making trajectory for spontaneous pneumothorax is shown in **Figure 2**. Since this patient presents clinical reasoning which strongly mimics acute pulmonary embolism (PE), such as a history of a long-distance flight and lower-leg discomfort, the embedding position of the initial state *r*_0_ is heavily biased toward PE.

**Figure 2:**
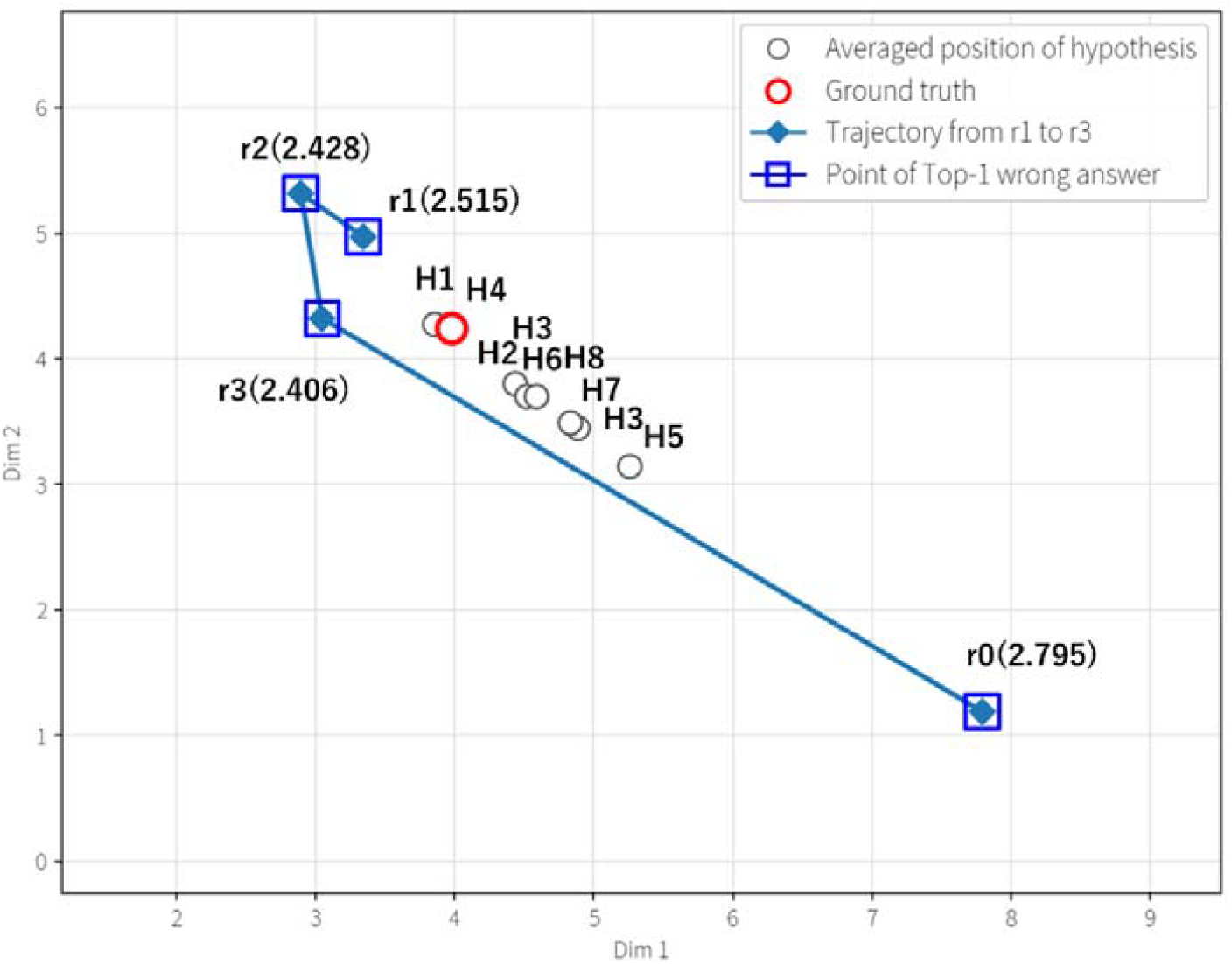
ISOMAP for inference trajectory for the ground truth disease, pneumonia. Converged on an incorrect hypothesis closer than the ground truth hypothesis. Here, the white circles represent the hypothesis position, where H1 indicates pulmonary embolism, H1 indicates acute coronary syndrome, H2 indicates aortic aneurysm, and H3 indicates spontaneous pneumothorax, H4 indicates pneumonia, H5 indicates pericarditis, H6 indicates Pericarditis, H7 indicates lung tumor, H8 indicates unstable seizures Other symbols are the same as in Figure 1.

Although diagnostic pseudo-tests such as CT pulmonary angiography in *r*_1_ returned negative or ambiguous findings, the initial baseline system lacked mathematically defined rules for explicit repulsive forces (repulsive vectors) to competing hypotheses exerted by negative findings, as well as boundary-value handling rules. Consequently, the plot upon updating at *r*_l_ could not form a clear update vector toward the target of spontaneous pneumothorax, resulting in stagnation and wandering within the space.

From *r*_2_ to the final round *r*_3_, the inference plot (blue diamond) deviates completely from spontaneous pneumothorax geometrically. Deu to the existence of incorrect competing attractor PE, it results as a stagnation and mis-convergence to the Top-1 incorrect answer points (blue squares). This reflects a fundamental structural error of the baseline system, such that negative exclusion can not be implemented in the probability space, and it then demonstrates a typical wandering dynamic where it converges on an incorrect hypothesis rather than the correct hypothesis.

In this case, Shannon entropy is not stable even at final round, its value 2.4 remains relatively high, in complete contrast with case A. The wandering dynamic of the inference state vector represents the non-convergence of Shannon entropy (**Table 2 and Figure 2**).

**Table 2:** Showing probability and entropy after each round, as shown in Table 1, in case B.

| Round | $p_{H1}$ | $p_{H2}$ | $p_{H3}$ | $p_{H4}$ | $p_{H5}$ | $p_{H6}$ | $p_{H7}$ | $p_{H8}$ | Entropy |
| --- | --- | --- | --- | --- | --- | --- | --- | --- | --- |
| $r_0$ | 0.222 | 0.194 | 0.167 | 0.139 | 0.111 | 0.083 | 0.056 | 0.028 | 2.795 |
| $r_1$ | 0.38 | 0.18 | 0.1 | 0.155 | 0.05 | 0.08 | 0.035 | 0.02 | 2.515 |
|  | 0.39 | 0.15 | 0.09 | 0.22 | 0.04 | 0.06 | 0.03 | 0.02 | 2.428 |
|  | 0.38 | 0.16 | 0.08 | 0.24 | 0.04 | 0.05 | 0.03 | 0.02 | 2.406 |

### 5.3. Case C: Acute Cholangitis in a Clean Environment

- **Patient Profile:** 72-year-old male
- **Chief Complaint:** Fever, right upper quadrant pain, and jaundice
- **History of Present Illness:** Onset of right upper quadrant pain 2 days prior, followed by a fever in the 38°C range starting yesterday. The patient was brought to the emergency department after family members noticed scleral icterus. Associated symptoms included right hypochondrial pain, chills, and anorexia. No vomiting or diarrhea was reported. Dark urine was observed. The patient was alert and oriented but appeared acutely ill and fatigued.
- **Past Medical History:** History of cholelithiasis; hypertension
- **Medications / Allergies:** Antihypertensives; no known drug allergies
- **Social / Family History:** Mild alcohol consumption; non-smoker. Unremarkable family history.
- **Vital Signs and Physical Examination:** Body Temperature: 38.6°C, Heart Rate: 112 bpm, Blood Pressure: 96/60 mmHg, Respiratory Rate: 22 breaths/min, SpO_2_ : 96% (room air), Japan Coma Scale (JCS): 0. Physical examination revealed overt jaundice and tenderness in the right hypochondrium (right upper quadrant).

As an example of a structural error where the prioritization of clinical safety (severity triage) led to a drop in the disease hypothesis ranking, the trajectory of acute cholangitis in a clean environment in which pseudo-test presents either positive or negative result is shown in **Figure 3**. Tracing the inference state vector (blue diamonds) reveals that, from the initial state *r*_O_ (lower-right point) through *r*_l_, *r*_2_, and *r*_3_, the vector moves linearly along the shortest distance toward a position of the barycenter of acute cholangitis without any wandering or detours. Looking solely at the geometric positional relationship, at the final round (*r*_3_) state vector completely reaches the immediate vicinity of the ground truth (red circle), regarding an accurate approach.

**Figure 3:**
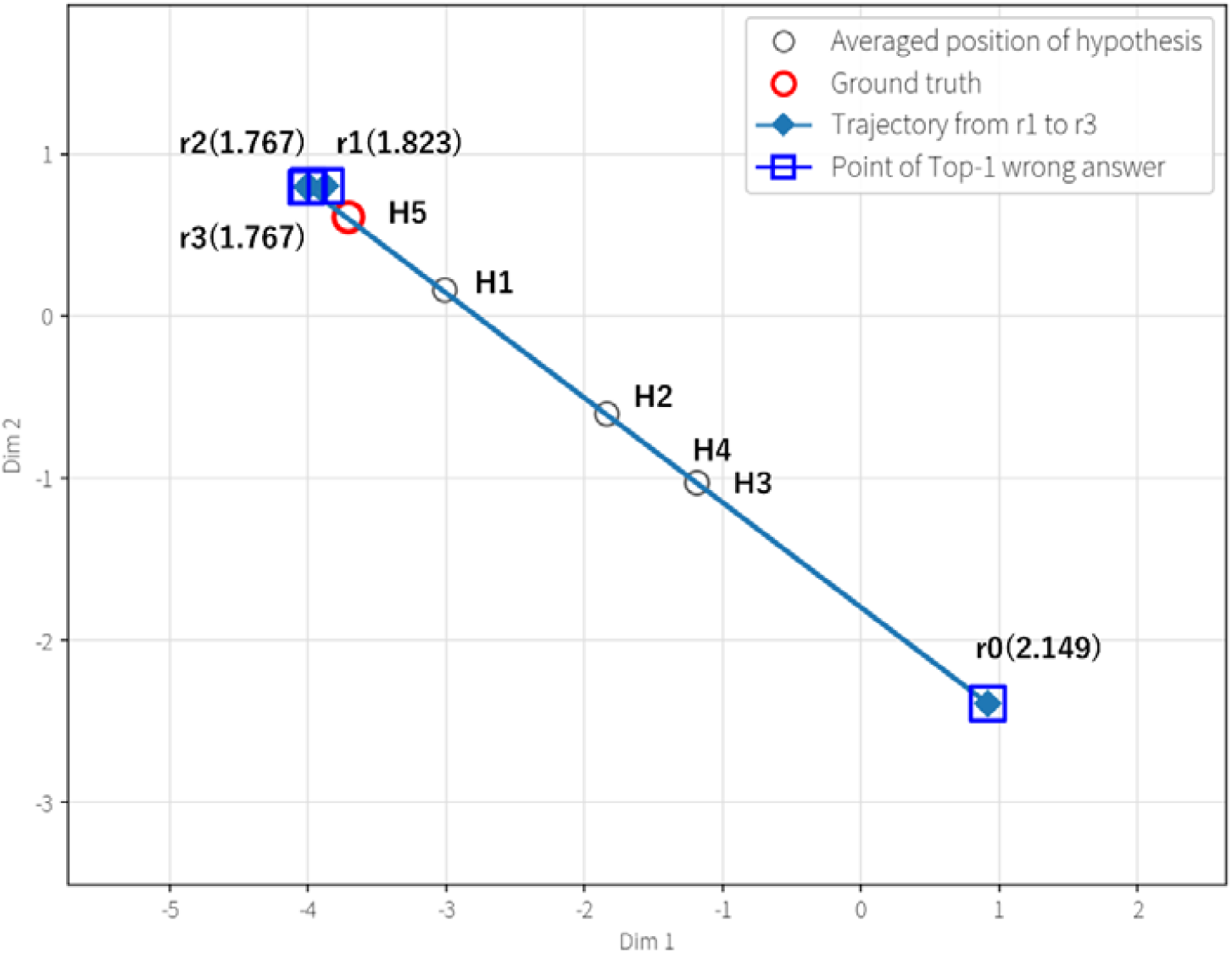
ISOMAP trajectory for the ground truth disease, Acute cholangitis. Converged to a position close to the ground truth hypothesis, but sepsis, which is clinically highly severe, was selected. Here, the white circles represent the hypothesis locations (H), where H1 indicates sepsis, H2 indicates acute cholecystitis, H3 indicates acute pancreatitis, H4 indicates acute hepatitis, and H5 indicates acute cholangitis. Other symbols are the same as in Figure 1.

However, those state vectors are enclosed by “Top-1 incorrect answer points” (blue squares) across all update steps. Clinically, sepsis, a severe systemic shock state occurring secondarily to the aggravation of acute cholangitis, is a critical red-flag condition that must be addressed with the highest priority in emergency care. In the baseline system, this physiological acute severity condition of sepsis and its anatomical etiology of acute cholangitis are conflated and output is the same ranking of hypothesis.

As a result, because the LLM prioritized clinical safety and assigned sepsis to the Top-1 position in its final output, the ranking of the true disease hypothesis remained in second place, even though geometrically it had reached right in front of the barycenter of the true cause, acute cholangitis. This reflects a state where clinical safety triage and evidence-based etiology identification undergo unjustified conflation and stagnation within the same evaluation space, representing a typical structural bottleneck that compromised Top-1 accuracy despite having converged closest to the ground-truth hypothesis.

In this case, the behavior of convergence of Shannon entropy is similar to the case A, although the Top-1 ranking of hypothesis is incorrect. This issue is also same as the typical bottleneck of Top-1 accuracy (**Table 3 and Figure 3**).

**Table 3:** Showing probability and entropy after each round, as shown in Table 1, in case C.

| Round | $p_{H1}$ | $p_{H2}$ | $p_{H3}$ | $p_{H4}$ | $p_{H5}$ | Entropy |
| --- | --- | --- | --- | --- | --- | --- |
| $r_0$ | 0.333 | 0.2667 | 0.2 | 0.1333 | 0.0667 | 2.149 |
|  | 0.43 | 0.12 | 0.05 | 0.04 | 0.36 | 1.823 |
|  | 0.45 | 0.11 | 0.05 | 0.03 | 0.36 | 1.767 |
|  | 0.45 | 0.11 | 0.05 | 0.03 | 0.36 | 1.767 |

## 6. Discussion

### 6.1. Proof of Concept: Validity of the Framework Through Pilot Evaluation

This study introduces a novel closed-loop, multi-agent clinical reasoning architecture, validated through a detailed geometric and information-theoretic analysis. Although the current experimental scope is deliberately confined to evaluating a baseline system via a pilot cohort of three representative emergency scenarios (the cases shown in Figures 1, 2, and 3), this focused evaluation validates the underlying methodology of visualizing epistemic uncertainty. By isolating these specific archetypes, a clean convergence (Figure 1), a noise-induced confounding trap (Figure 2), and a triage-etiology conflict (Figure 3), we successfully visualized the precise mathematical and topological mechanisms by which LLMs succeed or fail. Rather than relying on static, black-box accuracy metrics, this pilot trial demonstrates that coupling Shannon entropy with ISOMAP provides a deterministic, auditable blueprint of the AI’s cognitive momentum.

### 6.2 Superiority and Novelty Compared to Prior Visualization Paradigms

The novelty of this research lies in its capacity to resolve the critical geometric pathologies inherent in existing clinical reasoning visualization frameworks. Previous paradigms, such as standard Latent Space Trajectories on medical manifolds (22-24), suffer from metric distortion and topological stagnation. In those models, when a reasoning trajectory stalls, it is mathematically impossible to distinguish whether the model is exhibiting healthy clinical deliberation or experiencing an ungrounded epistemic freeze. Similarly, Concept Activation Vectors (25-27) and standard Concept Bottleneck Models (28,29) frequently result in epistemic occlusion, masking the model’s internal hesitation behind a falsely confident final probability output.

Our framework transcends these limitations through geometric-information coupling. By integrating the continuous rate of entropy decay with Barycentric ISOMAP embeddings, we assign a quantifiable thermodynamic energy state to the reasoning trajectory. As demonstrated in our baseline results, when the LLM in the case shown in Figure 2 was misled by pulmonary embolism confounders, our visualization did not merely show a wrong answer; it geometrically exposed the model being trapped in a high-entropy local minimum. By mathematically capturing both the directional alignment and the epistemic uncertainty simultaneously, this study achieves a level of transparent auditability that previous post-hoc explainability methods could not attain.

### 6.3 Real-World Clinical Significance

The integration of this visualization framework into real-world clinical environments holds transformative potential for mitigating automation bias and the deskilling of medical professionals (6,10). In acute emergency settings, clinicians operating under severe time constraints and cognitive fatigue are susceptible to premature closure, uncritically accepting algorithmic outputs (7-9).

Our visualization translates the abstract, black-box computations of an LLM into an intuitive, navigable 2D map. When a clinician observes a trajectory oscillating wildly with high entropy or getting trapped in a non-target cluster, it acts as an immediate, objective warning that the AI’s predictive distribution has degraded into high-variance stochasticity or algorithmic stagnation, prompting the clinician to override the system and exercise independent judgment. Conversely, a clean, low-entropy trajectory converging directly onto a diagnostic node validates the AI’s reasoning path. By rendering the machine’s internal confusion visible before the final hypothesis crystallizes, this system dynamically calibrates physician trust, fostering a collaborative human-AI co-regulation that enhances diagnostic safety rather than eroding human expertise.

### 6.4 Hypotheses for Algorithmic Improvement and Future Architecture Scaling

Because the current pilot study focused on auditing and visualizing the baseline LLM architecture, the structural reasoning errors (mis-convergence and stagnation) remained unresolved within the evaluation. However, by mathematically exposing the exact locations and causes of these cognitive failures on the latent manifold, this framework provides a blueprint for designing the next generation of clinical AI. Based on the geometric failures identified in this baseline trial, we propose the following architectural intervention hypotheses for future research:

#### Hypothesis 1: Decoupling Safety from Etiology via Dual-Channel Architecture (Addressing the case C in Figure 3)

The baseline system’s stagnation in the case shown in Figure 3 revealed that LLMs inherently conflate acute physiological threats (e.g., sepsis) with anatomical etiologies (e.g., acute cholangitis) within a single probability space. We hypothesize that implementing a dual-channel architecture—separating a Critical Safety Gate from the Etiology Ranking—will mathematically eliminate this triage-etiology conflation, allowing the model to accurately achieve Top-1 diagnosis without compromising life-saving safety alerts.

#### Hypothesis 2: Amplifying Repulsive Vectors for Negative Evidence (Addressing the case B in Figure 2)

The baseline model failed to rule out pulmonary embolism because it lacked structural sensitivity to negative or borderline test results. We hypothesize that introducing explicit, non-linear repulsive weights into the log-linear Bayesian Expected Information Gain (EIG) approximation will forcefully repel the trajectory away from competing attractors upon receiving negative clinical findings, preventing local minimum traps even in highly noisy clinical environments.

#### Hypothesis 3: Redundancy-Penalized Optimization for Clinical Efficiency

In clinical settings, minimizing unnecessary testing is paramount. We hypothesize that integrating a Redundancy-Penalized EIG function, which dynamically penalizes candidate tests that share overlapping information channels with previously executed tests, into the agent’s decision-making logic will drastically reduce the required Minimum Test Set (MTS), optimizing both healthcare costs and patient burden while maintaining diagnostic precision.

### 6.5 Future Research Proposal

To transition this framework from a conceptual pilot to a generalized clinical standard, future research must implement the proposed architectural interventions and scale the evaluation across diverse, large-scale clinical datasets. First, we propose conducting retrospective validations using massive, multi-center Electronic Health Record databases (e.g., MIMIC-IV) to test the robustness of the Barycentric ISOMAP trajectories across tens of thousands of heterogeneous clinical presentations.

Second, the pseudo-likelihood estimations generated by the LLM within the EIG approximation must be rigorously calibrated against real-world epidemiological statistics and expert physician consensus using modified Delphi methods. Finally, we propose prospective, randomized clinical trials comparing the diagnostic accuracy and automation bias rates of physicians using standard LLM outputs versus those utilizing our real-time, entropy-coupled trajectory visualizations. Such comprehensive scaling will firmly establish this architecture as the foundational blueprint for safe, transparent, and cognitively synergistic clinical AI.

## 7. Conclusion

This pilot study provides a novel, information-theoretically grounded framework for visualizing and auditing dynamic epistemic uncertainty in clinical LLM reasoning. By coupling predictive Shannon entropy with Barycentric ISOMAP latent space embeddings, our approach transcends static, black-box outcome metrics, offering an intuitive, two-dimensional blueprint of an AI agent’s internal cognitive momentum.

Through the geometric trajectory analysis of three representative emergency scenarios, this research successfully exposed the precise topological pathologies underlying baseline LLM reasoning failures, specifically, mis-convergence trapped by confounding noise attractors and structural stagnation driven by triage-etiology conflation, while confirming smooth, low-entropy convergence along geodesic paths in unconfounded regimes.

By rendering machine hesitation and cognitive divergence visible prior to final hypothesis crystallization, this framework provides an essential mechanism for dynamic trust calibration and human-AI co-regulation in high-stakes clinical settings. Crucially, the structural bottlenecks identified in this trial lay down a clear mathematical rationale for future architectural interventions, including dual-channel safety decoupling and repulsive negative-evidence weighting. Implementing these algorithmic enhancements and scaling evaluation across large-scale Electronic Health Record databases and prospective clinical trials will firmly establish this architecture as a foundational blueprint for safe, transparent, and synergistic clinical artificial intelligence.

## Data Availability

All data produced in the present study are available upon reasonable request to the authors

## References

1. Thirunavukarasu AJ, Ting, DSJ, Elangovan K. et al. Large language models in medicine. Nat Med 29, 1930–1940 (2023). doi:10.1038/s41591-023-02448-8

2. Liu F., Zhou H., Gu B. et al. Application of large language models in medicine. Nat Rev Bioeng 3, 445–464 (2025). doi:10.1038/s44222-025-00279-5

3. Shah NH, Entwistle D, Pfeffer MA. Creation and Adoption of Large Language Models in Medicine. JAMA. 2023;330(9):866–869. doi:10.1001/jama.2023.14217

4. Heudel PE, Crochet H, Filori Q, Bachelot T, Blay JY. Artificial intelligence in medicine: a scoping review of the risk of deskilling and loss of expertise among physicians. ESMO Real World Data Digit Oncol. 2026;12:100693. Published 2026 Mar 19. doi:10.1016/j.esmorw.2026.100693

5. El Tarhouny S, Farghaly A. Deskilling dilemma: brain over automation. Front Med (Lausanne). 2026;13:1765692. Published 2026 Feb 3. doi:10.3389/fmed.2026.1765692

6. Wolters Kluwer. Patients, Doctors, and Nurses on AI: Similar Tools, Different Pathways, One Destination. Wolters Kluwer; 2026. Accessed July 29, 2026. https://assets.contenthub.wolterskluwer.com/api/public/content/3333130-2026-future-ready-healthcare-survey-report-pdf--774e854545?v=e4d07570

7. Rosbach E, Ganz J, Ammeling J, et al. Automation Bias in AI-Assisted Medical Decision-Making under Time Pressure in Computational Pathology. arXiv [Preprint]. Published November 1, 2024. Accessed July 29, 2026. doi:10.48550/arXiv.2411.00998

8. Qazi I, Ali A, Khawaja AU, et al. Automation Bias in Large Language Model–Assisted Diagnostic Reasoning among Physicians Trained in AI Literacy: A Randomized Clinical Trial. NEJM AI. 2026;3(5). doi:10.1056/AIoa2501001.

9. Mahajan A, Obermeyer Z, Daneshjou R, et al. Cognitive bias in clinical large language models. NPJ Digit Med. 2025;8:428. doi:10.1038/s41746-025-01790-0.

10. Saadeh MI, Janhonen J, Beer E, et al. Automation complacency: risks of abdicating medical decision making. AI Ethics. 2025;5(6):5783–5793. doi:10.1007/s43681-025-00825-2.

11. Challen R, Denny J, Pitt M, et al. Artificial intelligence, bias and clinical safety. BMJ Quality & Safety. 2019;28:231–237. doi:10.1136/bmjqs-2018-008370.

12. Budzyń K, Romańczyk M, Kitala D, et al. Endoscopist deskilling risk after exposure to artificial intelligence in colonoscopy: a multicentre, observational study. Lancet Gastroenterol Hepatol. 2025;10(10):896–896. doi:10.1016/S2468-1253(25)00133-5.

13. Logg JM, Minson JA, Moore DA. Algorithm appreciation: people prefer algorithmic to human judgment. Organ Behav Hum Decis Process. 2019;151:90–103. doi:10.1016/j.obhdp.2018.12.005.

14. Al-Anezi FM. Generative artificial intelligence in healthcare: automation bias, deskilling, and cognitive implications—a systematic review. J Healthc Leadersh. 2026;18:590498. doi:10.2147/JHL.S590498.

15. Goddard K, Roudsari A, Wyatt JC. Automation bias: a systematic review of frequency, effect mediators, and mitigators. J Am Med Inform Assoc. 2012;19(1):121–127. doi:10.1136/amiajnl-2011-000089

16. Jorritsma W, Cnossen F, van Ooijen PMA. Improving the radiologist-CAD interaction: designing for appropriate trust. Clin Radiol. 2015;70(2):115–115. doi:10.1016/j.crad.2014.09.017.

17. Patterson F, Kunar MA. The message matters: changes to binary Computer Aided Detection recommendations affect cancer detection in low prevalence search. Cogn Res Princ Implic. 2024;9(1):59. Published 2024 Sep 2. doi:10.1186/s41235-024-00576-4

18. Lee MH. From accuracy to readiness: metrics and benchmarks for human-AI decision-making. In: Extended Abstracts of the 2026 CHI Conference on Human Factors in Computing Systems (CHI EA ‘26). Association for Computing Machinery; 2026:1–10. doi:10.1145/3772363.3798377.

19. Ibrahim L, Collins KM, Kim SSY, et al. Measuring and mitigating overreliance to build human-compatible AI. arXiv [Preprint]. Published May 20, 2026. doi:10.48550/arXiv.2509.08010.

20. Ng DTK, Leung JKL, Chu SKW, Qiao MS. Conceptualizing AI literacy: an exploratory review. Comput Educ Artif Intell. 2021;2:100041. doi:10.1016/j.caeai.2021.100041.

21. Ryoo Y, Bakpayev M, Jeon YA, Kim K, Yoon S. High hopes, hard falls: consumer expectations and reactions to AI-human collaboration in advertising. Int J Advert. 2026;45(1):48–48. doi:10.1080/02650487.2025.2458996.

22. Patel S. The latent space hypothesis: toward universal medical representation learning. arXiv [Preprint]. Published June 4, 2025. doi:10.48550/arXiv.2506.04515.

23. Zamora-Resendiz R, Khurram I, Crivelli S. Towards maps of disease progression: biomedical large language model latent spaces for representing disease phenotypes and pseudotime. medRxiv [Preprint]. Published June 16, 2024. Updated July 8, 2024. doi:10.1101/2024.06.16.24308979.

24. Chen Q, Wang X, Chen R, et al. A unified AI-driven multimodal framework integrating visual sensing and wearable sensors for robust human motion monitoring in biomedical applications. Sensors (Basel). 2026;26(8):2314. doi:10.3390/s26082314.

25. Nicolson A, Schut L, Noble JA, Gal Y. Explaining explainability: understanding concept activation vectors. arXiv [Preprint]. Published April 4, 2024. doi:10.48550/arXiv.2404.03713.

26. Kim B, Wattenberg M, Gilmer J, et al. Interpretability beyond feature attribution: quantitative testing with concept activation vectors (TCAV). In: Proceedings of the 35th International Conference on Machine Learning. Proceedings of Machine Learning Research. 2018;80:2668–2677.

27. Brenner A, Knispel F, Fischer FP, et al. Concept-based AI interpretability in physiological time-series data: example of abnormality detection in electroencephalography. Comput Methods Programs Biomed. 2024;257:108448. doi:10.1016/j.cmpb.2024.108448.

28. De Felice G, Casanova Flores A, De Santis F, et al. Causally reliable concept bottleneck models. arXiv [Preprint]. Published March 6, 2025. doi:10.48550/arXiv.2503.04363.

29. Pang W, Ke X, Tsutsui S, Wen B. Integrating clinical knowledge into concept bottleneck models. arXiv [Preprint]. Published July 9, 2024. doi:10.48550/arXiv.2407.06600.

30. Hu L, Ren C, Hu Z, et al. Editable concept bottleneck models. In: Proceedings of the 42nd International Conference on Machine Learning. Proceedings of Machine Learning Research. 2025;267:24678–24726.

